# A descriptive study of the impact of diseases control and prevention on the epidemics dynamics and clinical features of SARS-CoV-2 outbreak in Shanghai, lessons learned for metropolis epidemics prevention

**DOI:** 10.1101/2020.02.19.20025031

**Authors:** Hongzhou Lu, Jingwen Ai, Yinzhong Shen, Yang Li, Tao Li, Xian Zhou, Haocheng Zhang, Qiran Zhang, Yun Ling, Sheng Wang, Hongping Qu, Yuan Gao, Yingchuan Li, Kanglong Yu, Duming Zhu, Hecheng Zhu, Rui Tian, Mei Zeng, Qiang Li, Yuanlin Song, Xiangyang Li, Jinfu Xu, Jie Xu, Enqiang Mao, Bijie Hu, Xin Li, Lei Zhu, Wenhong Zhang, on behalf of the Shanghai Clinical Treatment Expert Group for SARS-CoV-2

## Abstract

**Objective:** To describe and evaluate the impact of diseases control and prevention on epidemics dynamics and clinical features of SARS-CoV-2 outbreak in Shanghai.

**Design:** A retrospective descriptive study

**Setting:** China

**Participants:** Epidemiology information was collected from publicly accessible database. 265 patients admitted to Shanghai Public Health Center with confirmed COVID-19 were enrolled for clinical features analysis.

**Main outcome measure:** Prevention and control measures taken by Shanghai government, epidemiological, demographic, clinical, laboratory and radiology data were collected. Weibull distribution, Chi-square test, Fisher’s exact test, t test or Mann-Whitney U test were used in statistical analysis.

**Results:** COVID-19 transmission rate within Shanghai had reduced over 99% than previous speculated, and the exponential growth has been stopped so far. Epidemic was characterized by the first stage mainly composed of imported cases and the second stage where >50% of cases were local. The incubation period was 6.4 (95% CI 5.3 to 7.6) days and the mean onset-admission interval was 5.5 days (95% CI, 5.1 to 5.9). Median time for COVID-19 progressed to severe diseases were 8.5 days (IQR: 4.8-11.0 days). By February 11^th^, proportion of patients being mild, moderate, severe and critically ill were 1.9%(5/265), 89.8%(238/265), 3.8%(10/265), 4.5%(12/265), respectively; 47 people in our cohort were discharged, and 1 patient died.

**Conclusion:** Strict controlling of the transmission rate at the early stage of an epidemic in metropolis can quickly prohibit the spread of the diseases. Controlling local clusters is the key to prevent outbreaks from imported cases. Most COVID-19 severe cases progressed within 14 days of disease onset. Multiple systemic laboratory abnormalities had been observed before significant respiratory dysfunction.

## Introduction

Since December 2019, a novel coronavirus, later named by WHO as SARS-CoV-2, emerged in Wuhan, China [1-3] and rapidly spread throughout Hubei province, with clustered cases reported globally [4-5]. Until February 19^th^ 2020, the total reported confirmed corona virus disease 2019 (COVID-19) cases have reached more than 72,000 within mainland China, and 888 cases in other 25 countries globally and the increasing number of cases and widening geographical spread have raised concerns internationally[6]. Up until now, multiple studies have described the clinical characteristics of the COVID-19, including fever, fatigue, cough etc. Various researches reported intensive care unit (ICU) admission rate between 19.1% to 32%[7-9], but this data mainly came from Hubei regions, and it might be biased towards detecting severe cases at the beginning of the epidemic. Thus, clinical features from regions outside Hubei can further assist the understanding of the disease’s epidemiological and clinical characteristics. Shanghai reported its first case of SARS-CoV-2 infection in January 20^th^ 2020, and the total confirmed cases have reached to 333 cases by February 19^th^. As one of the global metropolis, Shanghai faces a relatively higher epidemics danger due to the substantial population mobility, and the risk is even doubled as Chunyun(a huge population flows during the Spring Festival) collided with the epidemics. During this outbreak, Shanghai has issued a number of strict measures to lower the transmissibility, including the shutdown of all large entertainment venue, reducing passenger flow and very strong social propaganda. Therefore, whether the Shanghai model has an impact on the epidemics dynamics and disease control is an important reference to the other metropolises around the world. In our study, we aimed to retrospectively describe the impact of diseases control on epidemics dynamics and clinical features of SARS-CoV-2 outbreak in Shanghai, and to provide valuable experience for other metropolises around the world.

## Methods

### Study design and participants

This is a retrospective, single-center cohort study, recruiting all patients admitted to Shanghai Public Health Center (SHPHC) diagnosed with coronavirus disease 2019 (COVID 2019) according to WHO interim guidance before Feb 7^th^, 2020. According to the arrangement of government, almost all adult patients from whole Shanghai were admitted to SHPHC once coronavirus disease 2019(COVID-19) was confirmed by real-time PCR. The study was approved by SHPHC ethics committee and oral consent was obtained from patients.

All patients admitted to Shanghai Public Health Center diagnosed with coronavirus disease COVID-19 according to WHO interim guidance were enrolled in this study [10]. Two cohorts were generated in this study, mild-moderate cohort and severe-critically ill cohort. All COVID-19 patients are classified as mild to critically ill cases at admission, according to COVID-19 Guidelines (the fifth version) made by National Health Commission of the People’s Republic of China. The classifying criteria was as follows:

Mild: Presenting mild symptoms and normal radiology manifestation in both lungs. Moderate (typical): Presenting typical symptoms (fever, cough and other respiratory symptoms) and radiology manifestation suggesting pneumonia.

Severe: Presenting any one of the followings:

1. Respiratory distress, respiratory rates ≥30 per minute;
2. Pulse oxygen saturation ≤93% on room air
3. Oxygenation Index (PaO2/FiO2) ≤300mmHg

Critically ill: Presenting any one of the followings

1. Respiratory failure where invasive ventilation is necessary
2. Signs of shock (circulatory failure)
3. Failure of any other organ where ICU care is necessary

## Data Collection

The cumulative numbers of confirmed and suspected cases were collected from Shanghai CDC, which were updated daily and publicly accessible. We obtained epidemiological, demographic, clinical, laboratory and radiology data from patients’ medical records. The data were reviewed by a trained team of physicians. Information recorded included demographic data, medical history, exposure history, comorbidities, symptoms, laboratory findings at baseline and chest x-ray or computed tomographic (CT) scans. The date of disease onset was defined as the day when the symptom was noticed.

## Statistical analysis

The incubation period distribution (i.e., the time delay from suspected contact to illness onset), and the onset-to-admission distribution was estimated by fitting a Weibull distribution to data on suspected exposure and onset dates in a subset of cases with detailed information available. Categorical variables were described as frequencies and percentages, and compared by Chi-square test or Fisher’s exact test between two cohorts. Continuous variables were described as median and interquartile range (IQR) values.

Means for continuous variables were compared using independent group t-tests when the data were normally distributed; otherwise, the Mann-Whitney test was used. For comparisons, a 2-sided α of less than 0.05 was considered statistically significant. Results of laboratory tests were also standardized and arranged by unsupervised hierarchical clustering to identify the similarities, differences and characteristics, with Euclidean distance measure and average linkage between groups methods. Statistical analyses were performed using SPSS (Statistical Package for the Social Sciences) version 22.0 software (SPSS Inc) and R software. Analyses of the incubation period, extensive predictive analysis were performed with the use of MATLAB software (R2016b).

## Results

### Epidemiological analysis

As of February, 19^th^ 2020, a total number of 72,531 confirmed COVID-19 cases have been reported according to reports from 31 provinces (autonomous regions, municipalities) and the Xinjiang Production and Construction Corps in mainland China. However, the number of newly increased cases, especially in regions outside Hubei provinces, has gradually decreased since February 5^th^ with the peak value of 3887. The same decreased trend can be seen in the newly increased cases in Shanghai, with continuous declined growth rate observed since February 4^th^. The total confirmed cases number of COVID-19 cases was 333 as of February 19^th^ 2020, of which the percentage of COVID-19 cases without travel history to Hubei province gradually surpassed 50%, according to the official report (Figure 1-A). The increased trend of cases without travel history to Hubei Provence and the confirmed 45 clusters cases in Shanghai implied that the second-generation cases in Shanghai had appeared gradually. In our study, the earliest symptom onset of all confirmed patients can be traced back to January 6^th^ 2020. The development of the epidemic followed an exponential growth and a decline in newly reported cases (Figure 2).

**Figure 1:**
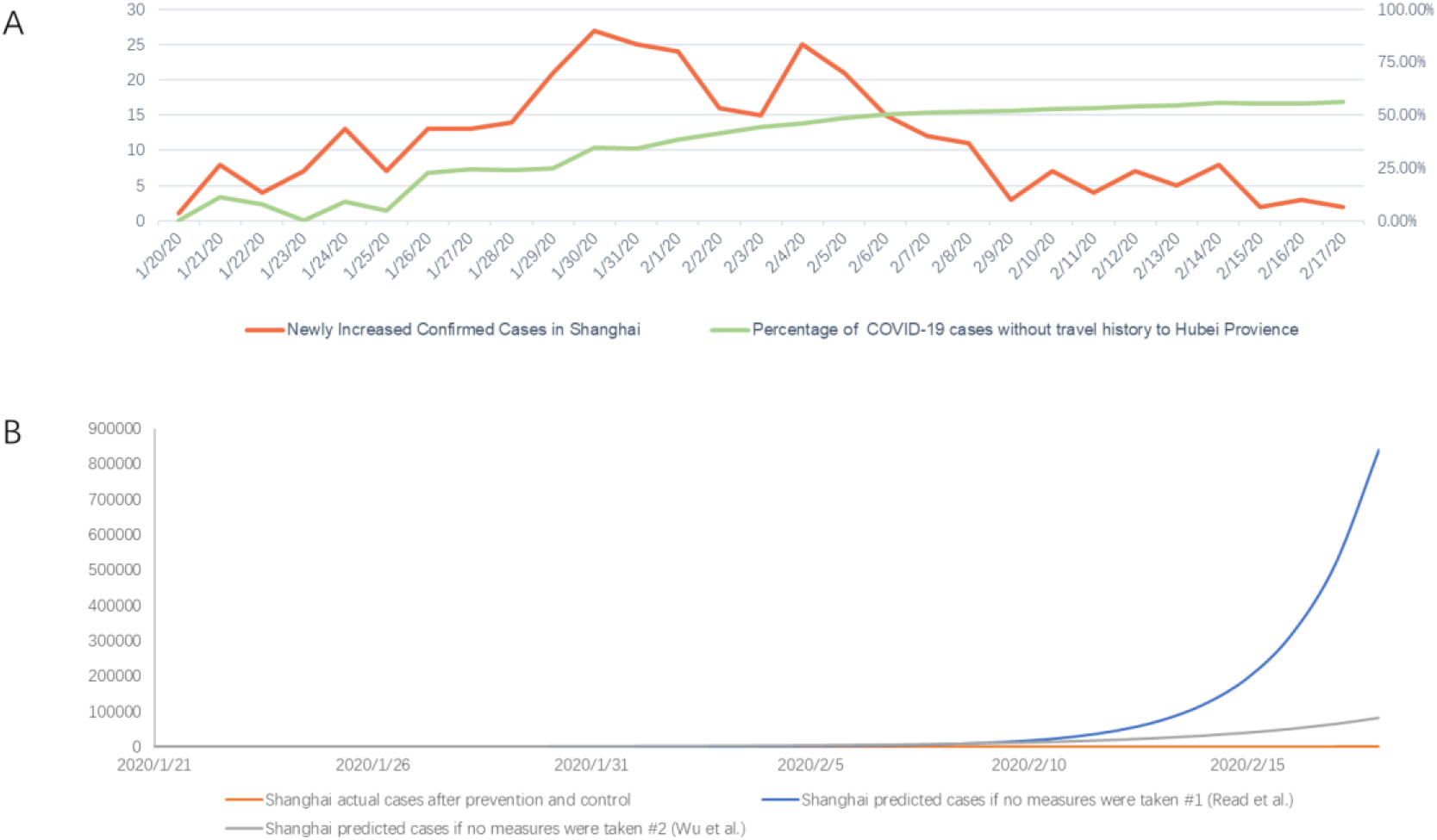
COVID-19 trends in Shanghai. (A) Newly increased confirmed cases in Shanghai and the percentage of COVID-19 cases without travel history to Hubei Province. (B) Actual and predicted COVID-19 cases in Shanghai.

**Figure 2.**
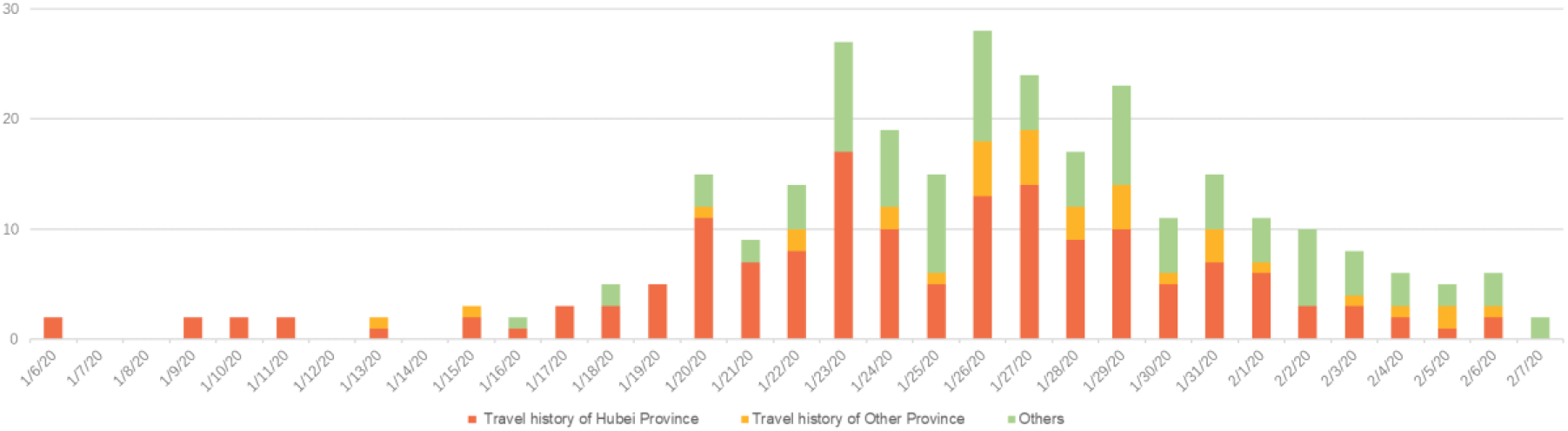
Onset of Illness among confirmed cases of COVID-19 in Shanghai, China. The date of disease onset was defined as the day when the symptom was noticed.

The study examined data on suspected exposures among all 265 laboratory-confirmed cases with detailed, 37 of whom had credible information on contacts to calculate the incubation period distribution. The mean incubation period is 6.4 days (95% CI 5.3 to 7.6) and the 5th and 95th percentile of the distribution was 0.97 and 13.10, respectively (Figure 3-A). The mean onset-admission interval was 5.5 days (95% CI, 5.1 to 5.9, SD 3.5). The 5th and 95th percentile of the distribution was 1 and 11.99, respectively (Figure 3-B).

**Figure 3.**
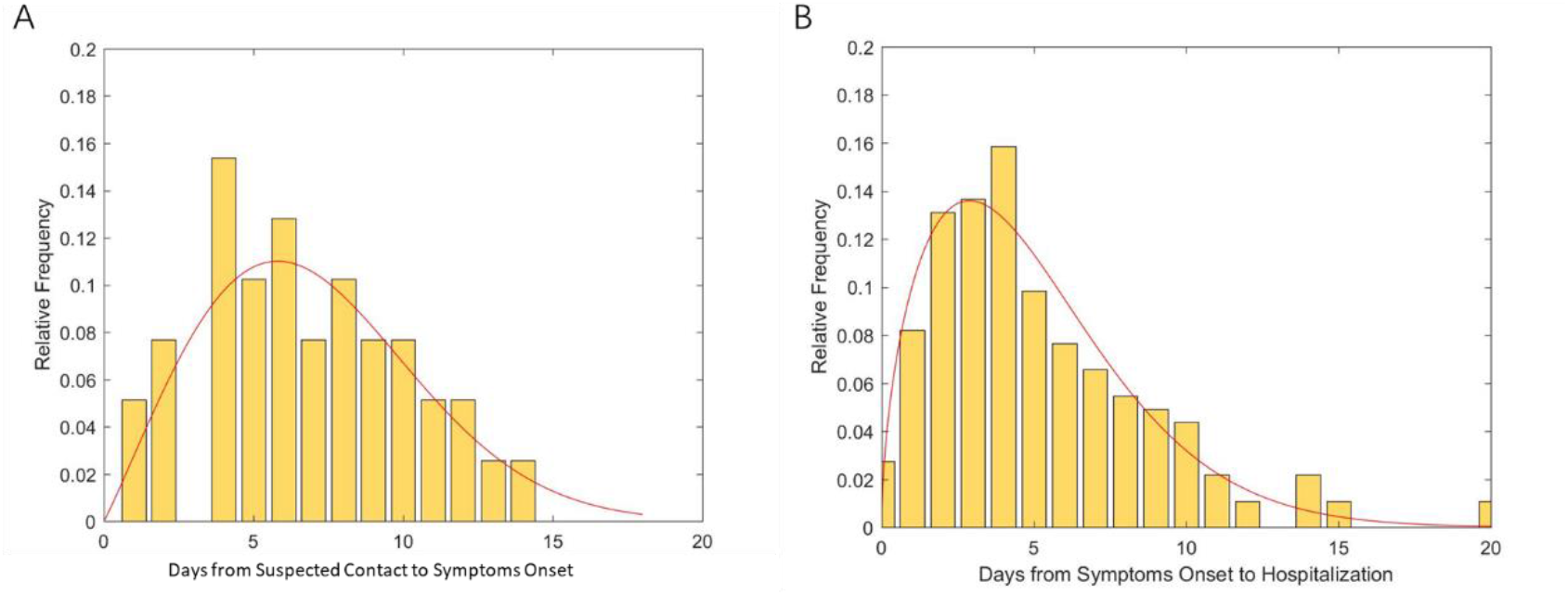
Key time-to-event distributions.

We performed an extensive analysis to February 9^th^ 2020 using the previously reported predictive parameterized transmission models in Shanghai and compared with the actual reported confirmed cases [11-12]. The previous predictive model was based on the hypothesis that no particular effective measures had been taken around the world and the transmission among population had not been controlled. Our result showed that the transmission rate within Shanghai had all decreased more than 95% than previously speculated, suggesting that the prevention and control interventions taken by Shanghai are of great impact on the overall control of the diseases during the current disease stage (Figure 1-B).

### Baseline clinical features

220 (90.9%) patients had fever before admission. Nearly half of patients were presented with pneumonia symptoms including cough (49.4%), expectoration (23.0%), chest pain (2.3%). Other common symptoms included fatigue (25.3%), inappetences (11.7%), headache (9.8%), myalgia (8.7%). Only 6.4% patients had diarrhea. Most symptom profile were comparable between mild-moderate patients (N=243) and severe-critically ill patients (N=22) while dyspnea occurred in a significantly higher proportion of severe or critical ill patients (1/237 vs. 4/22, p<0.001). Eleven patients were asymptomatic on admission (Table 1).

**Table 1.** Clinical symptoms, comorbidities, and radiology findings at admission.

|  | All Patients<br>N=265 | Mild-moderate<br>N=243 | Severe-critically<br>ill<br>N=22 | P value |
| --- | --- | --- | --- | --- |
| **Symptoms** |  |  |  |  |
| Fever | 220 (90.9%) | 200 (82.3%) | 20 (90.9%) | 0.303 |
| Cough | 131 (49.4%) | 124 (51.0%) | 7 (31.8%) | 0.084 |
| Expectoration | 61 (23.0%) | 55 (22.6%) | 6 (27.3%) | 0.621 |
| Fatigue | 67 (25.3%) | 60 (24.7%) | 7 (31.8%) | 0.461 |
| Inappetence | 31 (11.7%) | 28 (11.5%) | 3 (13.6%) | 0.730 |
| Headache | 26 (9.8%) | 25 (10.3%) | 1 (4.5%) | 0.707 |
| Myalgia | 23 (8.7%) | 21 (8.6%) | 2 (9.1%) | 0.999 |
| Chest tightness | 12 (4.5%) | 9 (3.7%) | 3 (13.6%) | 0.067 |
| Chest pain | 6 (2.3%) | 5 (2.1%) | 1 (4.5%) | 0.409 |
| Dyspnea | 5 (1.9%) | 1 (0.4%) | 4 (18.2%) | <0.0001 |
| Rhinorrhea | 16 (6.0%) | 16 (6.6%) | 0 (0.0%) | 0.376 |
| Sore throat | 12 (4.5%) | 12 (4.9%) | 0 (0.0%) | 0.607 |
| Diarrhea | 17 (6.4%) | 17 (7.0%) | 0 (0.0%) | 0.200 |
| Nausea or vomiting | 6 (2.3%) | 6 (2.5%) | 0 (0.0%) | 0.999 |
| **Comorbidities** |  |  |  |  |
| Hypertension | 52 (19.6%) | 42 (17.3%) | 10 (45.5%) | 0.004 |
| Diabetes Mellites | 21 (7.9%) | 15 (6.2%) | 6 (27.3%) | 0.004 |
| Coronary diseases | 14 (5.3%) | 10 (4.1%) | 4 (18.2%) | 0.021 |
| COPD | 4 (1.5%) | 2 (0.8%) | 2 (9.1%) | 0.036 |
| Tumor | 6 (2.3%) | 5 (2.1%) | 1 (4.5%) | 0.409 |
| Chronic renal diseases | 5 (1.9%) | 3 (1.2%) | 2 (9.1%) | 0.057 |
| Hyperlipidemia | 4 (1.5%) | 4 (1.6%) | 0 (0.0%) | 0.999 |
| Cerebrovascular diseases | 2 (0.8%) | 1 (0.4%) | 1 (4.5%) | 0.159 |
| Autoimmune diseases | 2 (0.8%) | 1 (0.4%) | 1 (4.5%) | 0.159 |
| Chronic liver diseases | 1 (0.4%) | 0 (0.0%) | 1 (4.5%) | 0.083 |
| **Radiology manifestation** |  |  |  | 0.263 |
| Normal | 9 (3.4%) | 9 (3.7%) | 0 (0.0%) |  |
| Unilateral involved | 51 (19.2%) | 49 (20.2%) | 2 (9.1%) |  |
| Bilateral involved | 205 (77.4%) | 185 (76.1%) | 20 (90.9%) |  |

Data were shown as n (%).
Abbreviation: COPD, chronic obstructive pulmonary disease

**Table 2.**
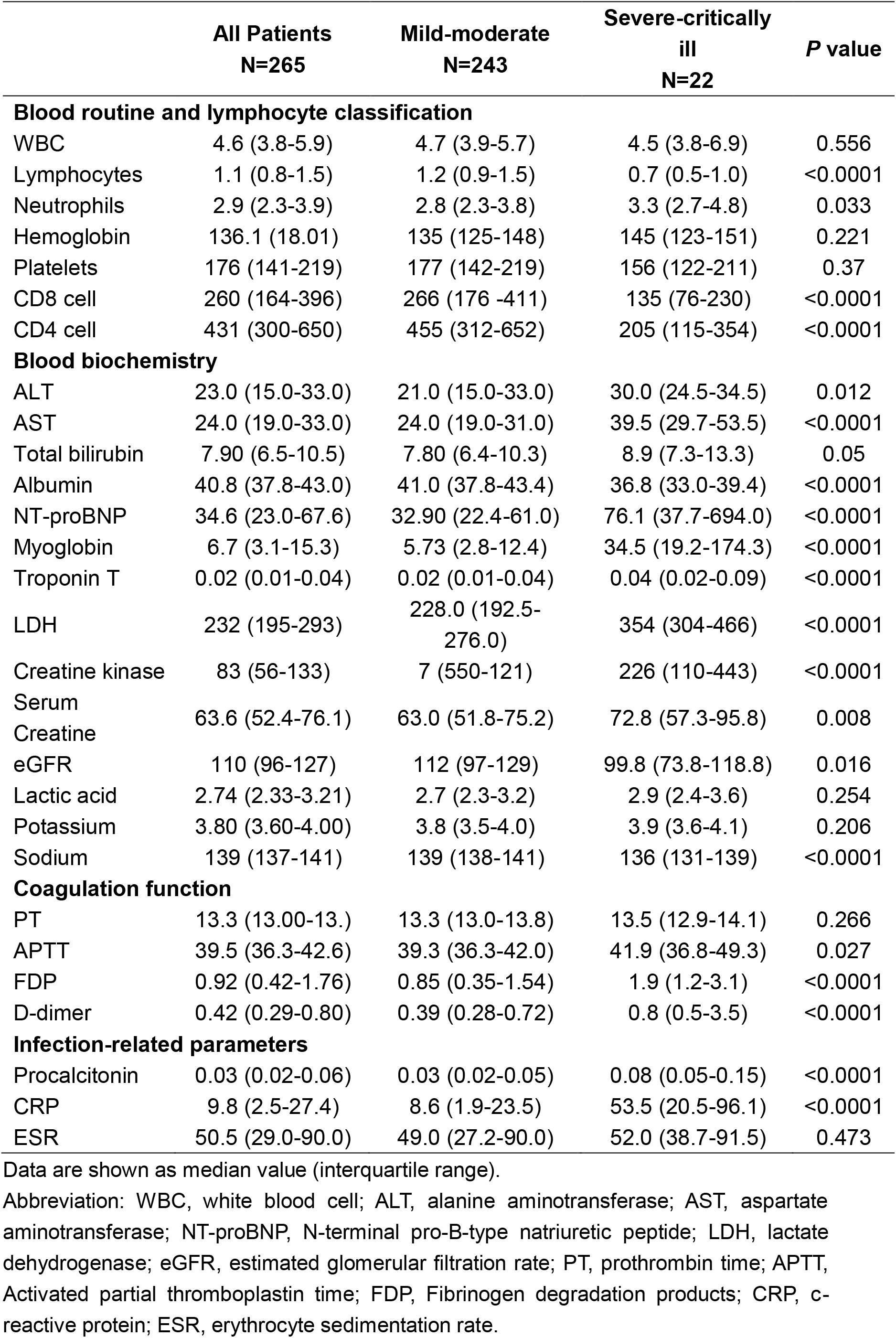
Laboratory examination at admission.

|  | All Patients<br>N=265 | Mild-moderate<br>N=243 | Severe-critically<br>ill<br>N=22 | P value |
| --- | --- | --- | --- | --- |
| **Blood routine and lymphocyte classification** |  |  |  |  |
| WBC | 4.6 (3.8-5.9) | 4.7 (3.9-5.7) | 4.5 (3.8-6.9) | 0.556 |
| Lymphocytes | 1.1 (0.8-1.5) | 1.2 (0.9-1.5) | 0.7 (0.5-1.0) | <0.0001 |
| Neutrophils | 2.9 (2.3-3.9) | 2.8 (2.3-3.8) | 3.3 (2.7-4.8) | 0.033 |
| Hemoglobin | 136.1 (18.01) | 135 (125-148) | 145 (123-151) | 0.221 |
| Platelets | 176 (141-219) | 177 (142-219) | 156 (122-211) | 0.37 |
| CD8 cell | 260 (164-396) | 266 (176 -411) | 135 (76-230) | <0.0001 |
| CD4 cell | 431 (300-650) | 455 (312-652) | 205 (115-354) | <0.0001 |
| **Blood biochemistry** |  |  |  |  |
| ALT | 23.0 (15.0-33.0) | 21.0 (15.0-33.0) | 30.0 (24.5-34.5) | 0.012 |
| AST | 24.0 (19.0-33.0) | 24.0 (19.0-31.0) | 39.5 (29.7-53.5) | <0.0001 |
| Total bilirubin | 7.90 (6.5-10.5) | 7.80 (6.4-10.3) | 8.9 (7.3-13.3) | 0.05 |
| Albumin | 40.8 (37.8-43.0) | 41.0 (37.8-43.4) | 36.8 (33.0-39.4) | <0.0001 |
| NT-proBNP | 34.6 (23.0-67.6) | 32.90 (22.4-61.0) | 76.1 (37.7-694.0) | <0.0001 |
| Myoglobin | 6.7 (3.1-15.3) | 5.73 (2.8-12.4) | 34.5 (19.2-174.3) | <0.0001 |
| Troponin T | 0.02 (0.01-0.04) | 0.02 (0.01-0.04) | 0.04 (0.02-0.09) | <0.0001 |
| LDH | 232 (195-293) | 228.0 (192.5-276.0) | 354 (304-466) | <0.0001 |
| Creatine kinase | 83 (56-133) | 7 (550-121) | 226 (110-443) | <0.0001 |
| Serum Creatine | 63.6 (52.4-76.1) | 63.0 (51.8-75.2) | 72.8 (57.3-95.8) | 0.008 |
| eGFR | 110 (96-127) | 112 (97-129) | 99.8 (73.8-118.8) | 0.016 |
| Lactic acid | 2.74 (2.33-3.21) | 2.7 (2.3-3.2) | 2.9 (2.4-3.6) | 0.254 |
| Potassium | 3.80 (3.60-4.00) | 3.8 (3.5-4.0) | 3.9 (3.6-4.1) | 0.206 |
| Sodium | 139 (137-141) | 139 (138-141) | 136 (131-139) | <0.0001 |
| **Coagulation function** |  |  |  |  |
| PT | 13.3 (13.00-13.) | 13.3 (13.0-13.8) | 13.5 (12.9-14.1) | 0.266 |
| APTT | 39.5 (36.3-42.6) | 39.3 (36.3-42.0) | 41.9 (36.8-49.3) | 0.027 |
| FDP | 0.92 (0.42-1.76) | 0.85 (0.35-1.54) | 1.9 (1.2-3.1) | <0.0001 |
| D-dimer | 0.42 (0.29-0.80) | 0.39 (0.28-0.72) | 0.8 (0.5-3.5) | <0.0001 |
| **Infection-related parameters** |  |  |  |  |
| Procalcitonin | 0.03 (0.02-0.06) | 0.03 (0.02-0.05) | 0.08 (0.05-0.15) | <0.0001 |
| CRP | 9.8 (2.5-27.4) | 8.6 (1.9-23.5) | 53.5 (20.5-96.1) | <0.0001 |
| ESR | 50.5 (29.0-90.0) | 49.0 (27.2-90.0) | 52.0 (38.7-91.5) | 0.473 |

Data are shown as median value (interquartile range).
Abbreviation: WBC, white blood cell; ALT, alanine aminotransferase; AST, aspartate aminotransferase; NT-proBNP, N-terminal pro-B-type natriuretic peptide; LDH, lactate dehydrogenase; eGFR, estimated glomerular filtration rate; PT, prothrombin time; APTT, Activated partial thromboplastin time; FDP, Fibrinogen degradation products; CRP, c-reactive protein; ESR, erythrocyte sedimentation rate.

All patients underwent chest X-ray or CT at admission (Table 2). 202 (76.2%) patients showed bilateral pneumonia while 51 (19.2%) patients showed unilateral pneumonia, and 9 (3.4%) patients showed almost no abnormalities. The most common abnormalities were multiple ground-glass opacities. All asymptomatic patients had findings that consistent with pneumonia.

On admission, 120 (45.3%) and 39 (14.7%) patients had lymphopenia and leukopenia respectively (Table 2). Most patients had normal levels of hemoglobin and platelets (92.5% and 82.0%, respectively). Elevated level of lactate dehydrogenase (LDH) and creatine kinase were detected in 106 (40.0%) and 46 (17.4%) cases, respectively. The elevation of erythrocyte sedimentation rate(ESR) and C-reactive protein (CRP) was common.

Compared with patients with mild and moderate COVID-19, those with severe or critically disease presented with extensively and significantly different laboratory parameters, including lymphocytes and neutrophils, myocardial zymogram (creatine kinase, myoglobin, troponin T, LDH, NT-proBNP), liver and renal function (alanine aminotransferase, aspartate aminotransferase, albumin, serum creatine, and eGFR), coagulation function (activated partial thromboplastin time, fibrin degradation products, and D-dimer), and infection-related biomarkers (CRP and procalcitonin). The counts of CD4+ and CD8+ cell of severe or critically ill patients were 205/μl and 135/μl, significantly lower than those of the mild or moderate patients (Figure 4).

**Figure 4.**
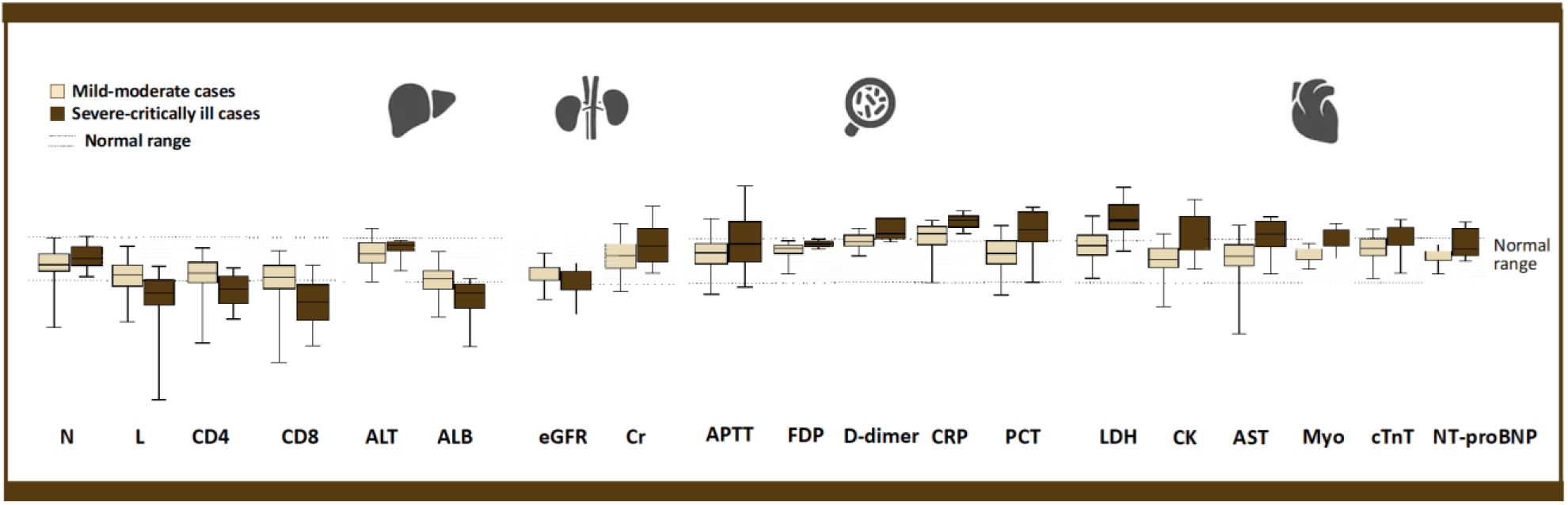
Significantly different laboratory parameters. Compared to the mild-moderate cohort, some laboratory results were obviously abnormal in Severe-critically ill cohort, suggesting impairments in different system in severe or critically ill COVID 2019 patients. Each box extends from the 25^th^ to 75^th^ percentiles, and each whisker goes down to the smallest value and up to the largest value (except for outliers. The shade between dotted line two dotted lines indicates the normal range of parameters. (The whiskers were generated using Tukey method, where values were regarded as outliers and not presented in this figure if greater than the sum of the 75^th^ percentile plus 1.5 IQR or less than 25^th^ percentile minus 1.5IQR. Boxes, whiskers and normal range change were extracted from graph of each parameter, adjusted to fit each other in fixed proportions, the distribution of the testing results and their relationship with normal range were not changed during the adjustment). All parameters presented in this figure had significant difference between two cohorts *(P*<0.05).

### Timeline and outcome of the disease progression

The proportion of patients with mild, moderate, severe and critically ill diseases on admission were 3.4% (9/265), 94.0% (249/265), 1.1% (3/265), and 1.5% (4/265), respectively. The spectrum of severity of diseases changed slightly as disease progressed. Of the 9 mild cases without pulmonary abnormality on admission, 4 showed bilateral or unilateral pneumonia in the subsequent chest CT tests which meant they were classified into moderate cases during hospitalization; and the remaining 5 had no changes in pulmonary imaging follow-ups for more than two weeks. While all the 11 asymptomatic patients had findings that consistent with pneumonia on routine CT examinations on admission. One of the patients was asymptomatic until she was discharged 11 days after being hospitalized. Two patients had subjective symptoms during the hospitalization. The remaining patients were still under observation in the hospital and had no symptoms. The median time for COVID-19 progressed to severe diseases was 8.5 days (IQR: 4.75-11.0 days), and to critically ill, requiring invasive mechanical ventilation, was 10.0 days (IQR: 5.5-11.0 days). There were 22 severely ill patients in the cohort, of which 21 were severely ill within 14 days of the course of the disease (Figure 5). When reevaluated the patients who were on day 14 of illness, we observed that 9.6% (17/177, 95%CI: 5.6-15.4%) had severe or critically ill COVID-19 while 86.7% (152/177) were stable, and other 4.5% (8/177) cured (Figure 6). By February 11^th^, the proportion of patients being mile, moderate, sever and critically ill were 1.9%(5/265), 89.8%(238/265), 3.8%(10/265), 4.5%(12/265), respectively; 47 (17.7%) people in our cohort were discharged, and 1 (0.38%) patient died. One of the critically ill patients was complicated with sepsis, two were treated with continuous renal replacement therapies, three patients were on extra-corporeal membrane oxygenation, and one patient recovered from severe disease to moderate condition.

**Figure 5.**
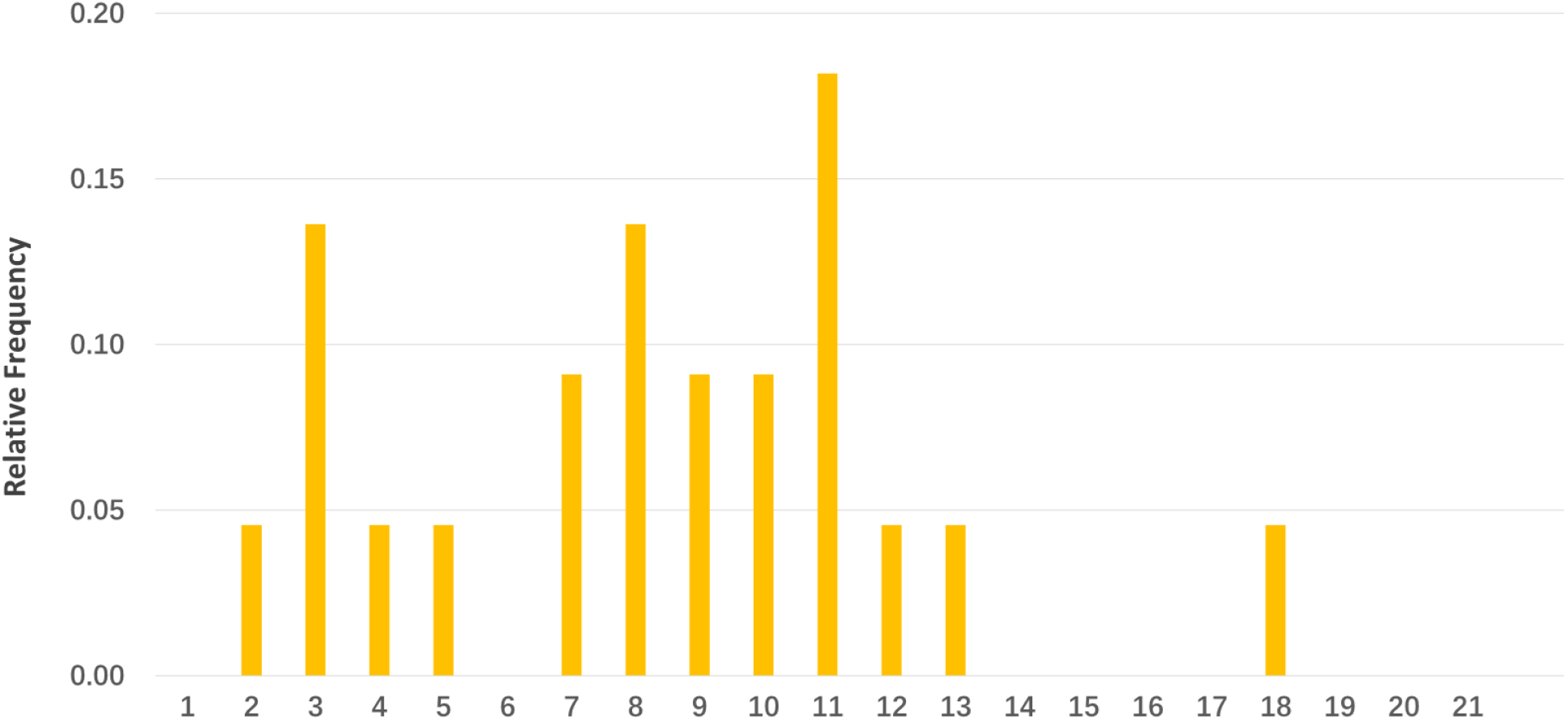
Days from illness onset to COVID-19 progression to severe illness. x-axis: the observation time from the onset of symptom for each patient. d0 is defined as the date diagnosis of COVID-19 confirmed.

**Figure 6.**
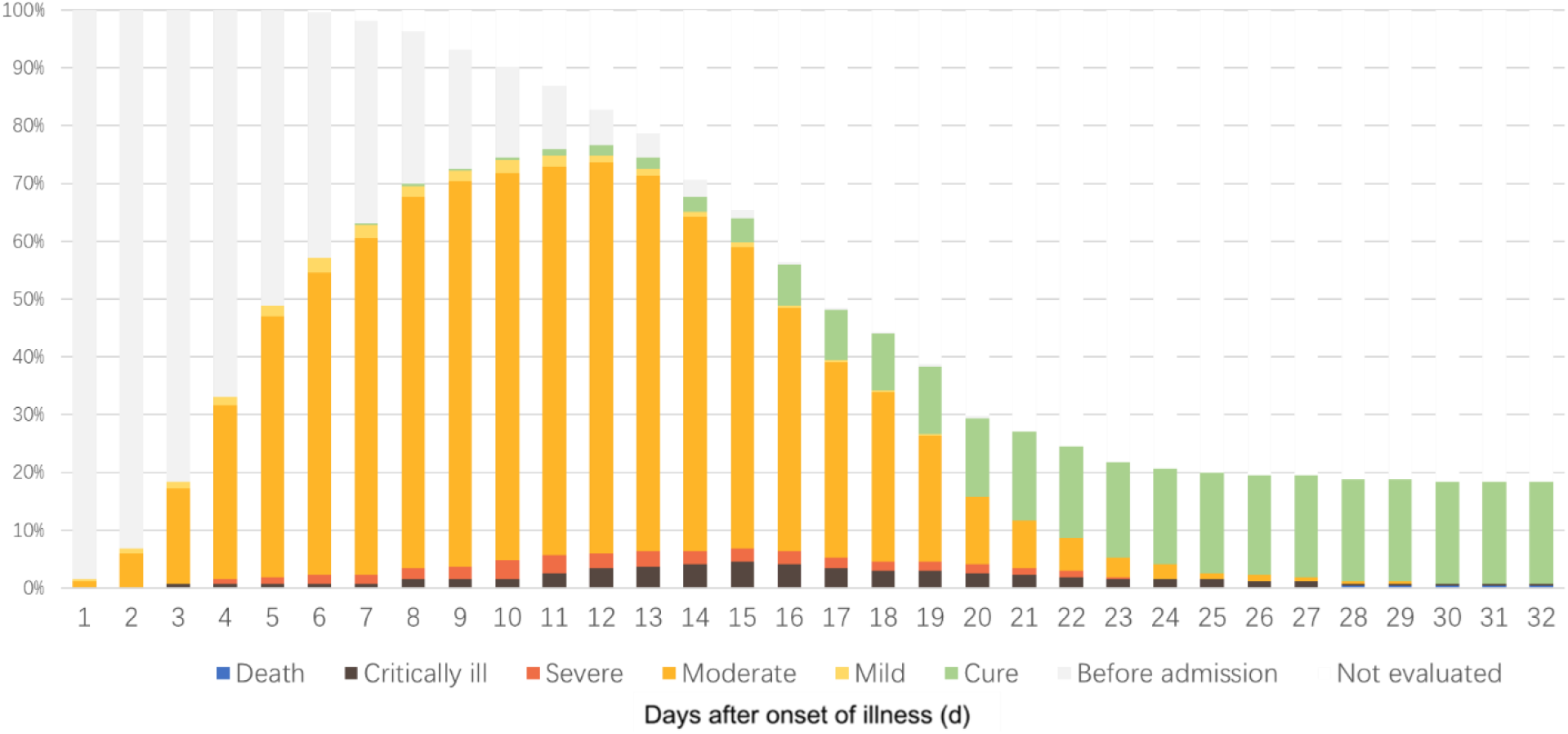
The patient composition and classification of the cohort. X-axis: the observation time from the onset of symptom for each patient. calculated; for asymptomatic patients, d0 is defined as the date diagnosis of COVID-19 confirmed. Y axis: the proportion of the entire cohort.

## Discussion

The first laboratory-confirmed case in Shanghai was reported in January 20^th^, and epidemic of Shanghai has experienced an increasing trend, during which the amplification of the newly diagnosed cases per day was increasing continuously. However, the trend had turned to a declining phase since January 31^st^. This phase was in accordance with the most regions outside Hubei province in China. In Hubei, newly diagnosed cases were still increasing in Hubei and on Feb 12^th^, because the clinical diagnosis was included in the guideline, the daily number of diagnosed patients had reached a spike of more than 10,000, allowing many suspected who haven’t the chance to be laboratory-confirmed to be admitted to the hospital for treatment. Therefore, in the near future, the local prevention and control of COVID-19 in Hubei would enter into a critical stage, where a turning point might be met if all measures were successively achieved.

Comparing the actual diseases trend in Shanghai with the previously estimated trends, we could find out that Shanghai exhibited a high-efficiency epidemics controlling ability and had stopped the exponential growth for now. The reason behind this is that during this epidemic, Chinese and Shanghai government has issued very strong and strict measures in restricting the transmission in local regions. The national transport department announced a 45% and 70.6% decline in the passenger flow at the beginning and the end of 2020 Spring Festival, respectively [13-14]. What’s more, Shanghai government cancelled all gathering activities and through propaganda of the officials, media and community, citizens were well-informed and well-educated of the importance of wearing face masks and washing hands frequently. Previous studies have suggested that the mobility reduction could not effectively lower the incidence rate while 50% transmissibility reduction could easily prevent the epidemics from further spreading [12]. In the case of Shanghai, the transmissibility was indeed greatly reduced, however, we believe the mobility reduction also played an indirect role in lowering the transmissibility. This is because that Chinese Spring Festival is characterized by hundreds of millions of people transporting from one region to another, and this massive gathering at the airports, train stations during transportation, if not controlled, would significantly increase the human to human transmission. Therefore, during this special period, controlling both mobility and transmissibility might be important. Other doubt lies in that previous studies have reported that the ascertainment rate in Wuhan is 5.1%[11], and suggested that other cities such as Shanghai, Beijing have the similar ascertainment rate, which would lead to our local case numbers fall below the true ones. However, in Shanghai for example, the suspected cases were approximately 20 folds of the laboratory-confirmed cases. Thus we expected the ascertainment rate outside Hubei province was closer to the actual epidemic size. But in the end, the disease prevention and control of the regions outside Hubei province would always heavily rely on the diminish of the epidemic in Hubei province, and the combination of Hubei epidemic control and strong measures to prevent local clustered cases outside Hubei will be the key to success.

In areas outside Hubei, the epidemics has transmitted from the first stage, in which imported cases composed of the main laboratory-confirmed cases, to the second stage, where imported cases and local sporadic or even clustered cases were simultaneously seen while imported cases from Hubei province gradually reduced due to the region shut down. In fact, clusters of cases within China and around the world without Hubei travelling history have been reported, indicating the shifting of the epidemic characteristics globally. In our study, we found that local cases increased approximately 5 days after the first the imported cases, possibly due to the incubation period after these local cases were infected. Also, the proportion of patients with a travel history to Hubei has gradually decreased, and the proportion of patients without a travel history to in Hubei has gradually increased. This suggests that strengthening prevention of local sporadic or clustered cases in areas outside Hubei in the future will be crucial for the second-stage epidemic control outside Hubei province.

In our study, the incubation period is similar as other studies previously reported, but the mean onset-admission interval was 5.478 days, shorter than 12.5 day initially reported in Wuhan[15], suggesting the timely control and prevention of the diseases spreading. One reason behind this is the improvement of the diagnostic ability of the disease, mainly the production of RT-PCR kit, which significantly shortened the diagnostic time frame.

The 11 asymptomatic patients in the Shanghai cohort were diagnosed after nucleic acid sampling of close contacts of confirmed patients. Routine chest CT examinations at admission revealed pneumonia in all of them. Therefore, these patients can only be considered asymptomatic but not in incubation. Although there was a reported case of incubation time up to 24 day [16], we believe that it is necessary to clarify whether an imaging assessment was performed before the onset of symptoms in this patient. However, it is worth noting that, at the epidemiological level, these asymptomatic patients are almost the same as those in the incubation period. A positive nasopharyngeal swab RT-PCR for COVID-19 in these patients suggests that they do have the potential to infect others. We therefore believe that more active inspections and evaluations of close contacts should be undertaken.

The proportion of severe and critically ill patients in Shanghai was significantly lower than that in Wuhan. We consider the proportion of patients with severe illness to be a very important indicator of the disease. First, regional mortality rate varies widely in the early stages of the disease outbreak because it is affected by many factors, such as life support equipment for critically ill patients and local medical conditions. In the absence of effective antiviral drugs, the proportion of critically ill patients depicts the natural course of the disease better than the mortality rate. Second, because the final clinical outcome of majority of the reported cases is typically unknown during a growing epidemic, dividing the cumulative reported deaths by reported cases will underestimate the mortality rate early in an epidemic[17]. We believe that the proportion of severe illness in Wuhan is biased towards detecting severe cases, partly because diagnostic capacity is limited at the start of an epidemic. Because of Shanghai’s active surveillance, especially for suspected cases with recent travel history to the affected region and close contacts, demonstrating by shorter onset-administration interval and 4% asymptomatic patients on admission, Shanghai should have picked up clinically milder cases as well as the more severe cases resulting a more reliable proportion of critically ill patients. To reduce the bias cause by mild case in early stage, we calculated the proportion of critically ill patients among those with an onset of 14 days or more, since more than 95% sever cases in Shanghai developed before that time. That proportion of 9.6% is substantially lower than SARS or MERS. The early clinical manifestations of COVID-19 patients in Shanghai were similar to those reported in Wuhan and elsewhere[7-9].

Our study showed relatively comprehensive laboratory test results in the published studies. Assessed through this systematic inspection, we can find that sever COVID-19 cases had extensive systemic laboratory abnormalities which indicated multisystem involvement had existed before significant respiratory abnormalities appeared. Previous studies have found that most patients had reduced lymphocytes and abnormal levels of many cytokines in critically ill patients [8,16]. Lymphocyte classification in our study showed that CD4 and CD8 cells both decreased in COVID-19 patients, and ∼15% patients had CD4 / CD8 <1. This may be related to a systemic inflammatory response caused by a cytokine storm, which is similar to that of SARS patients. The severity classification of COVID-19 patients in China mainly focuses on the respiratory system and oxygenation of patients. We believe more attention should be paid to other organs function of the patient in addition to the respiratory system. As follow-up continues, we can observe the outcome of more and more patients, and it should be possible to identify early risk factors that indicate a poor prognosis.

Our study has several limitations. First, as the epidemics has not ended yet, the effect of the control measures cannot be fully evaluated. Second, as most patients are still hospitalized at the time of manuscript submission, clinical outcome remains to be seen. Third, we did not measure cytokines and viral loads, which may be related to disease progression and severity.

## Conclusion

Strict measures on controlling disease transmissibility in a metropolis can quickly reduce the spread of a new infectious diseases to polar levels and stop its exponential growth in the short term. Controlling the local clusters is the key to prevent outbreaks due to imported cases. Our studies showed that the incubation period for COVID-19 is 6.438 days and 9.6% of COVID-19 cases were severe cases. The median time for COVID-19 progressing to severe diseases was 8.5 days and multiple systemic laboratory abnormalities which indicated multisystem involvement had been observed before significant respiratory abnormalities appeared.

## Data Availability

The data is available in the manuscript

## Consent from participate

Oral consent was obtained was obtained from the patients.

## Funding

This study was not funded.

## Competing interests

All authors declare no competing interests.

Acknowledgements

We acknowledge all health-care workers involved in the diagnosis and treatment of patients and show the greatest appreciation to all health workers for their valuable input to the control of diseases.

## Notes

### Competing Interest Statement

The authors have declared no competing interest.

